# Role of Walking Energetics and Perceived Fatigability on Mobility Differ by Walking Speed: The Study of Muscle, Mobility and Aging (SOMMA)

**DOI:** 10.1101/2023.11.05.23298138

**Authors:** Reagan E. Moffit, Terri Blackwell, Daniel E. Forman, Paul M. Coen, Barbara J. Nicklas, Yujia (Susanna) Qiao, Peggy M. Cawthon, Frederic G. S. Toledo, Bret H. Goodpaster, Steven R. Cummings, Anne B. Newman, Nancy W. Glynn

## Abstract

**Background:** Slow gait speed is a risk factor for poor health outcomes among older adults and may be driven by decreased energy availability and increased fatigability.

**Objective:** Examine walking energetics and perceived physical fatigability with gait speed among slower and faster walkers and understand whether fatigability statistically mediates the association between energetics and gait speed.

**Methods:** Perceived physical fatigability was assessed using the Pittsburgh Fatigability Scale (PFS) Physical score (range 0-50, higher=greater). A three-phase cardiopulmonary exercise treadmill test collected peak oxygen consumption (VO_2_peak mL/kg/min), energetic cost of walking per distance travelled (EC_W_, mL/kg/meter), and cost-capacity ratios (VO_2_/VO_2_peak, %). Gait speed was determined by 4m walk; slower (<1.01m/s) vs faster (≥1.01m/s) walkers were classified using median 4m gait speed. Linear regressions and statistical mediation analyses were conducted.

**Results:** Slower walkers had lower VO_2_peak, higher EC_W_ at preferred walking speed (PWS), and greater PFS Physical score compared to faster walkers (all p<0.05) (N=849). One standard-deviation higher increment of VO_2_peak, EC_W_ at PWS, cost-capacity ratios at PWS and slow walking speed (SWS), and PFS Physical score were associated with 0.1m/s faster (VO_2_peak only) or 0.02-0.09m/s slower gait speed. PFS Physical score was a significant statistical mediator in the associations between VO_2_peak (15.2%), cost-capacity ratio (15.9%), and EC_W_ at PWS (10.7%) with gait speed, and stronger among slower walkers.

**Conclusions:** Fitness and fatigability are associated with slower gait speed yet contributions may differ among slower and faster walkers. Future interventions may consider targeting fatigability among slower walkers and fitness among faster walkers.

## Introduction

Gait speed is a vital sign among older adults.[1,2] As adults age, both usual and fast gait speed decreases[3] and predicts poor health outcomes in older adults, including mortality,[4,5][6] mobility limitations,[7–9] difficulty completing activities of daily living (ADLs),[10] and increased healthcare cost and utilization.[11] Understanding factors associated with age-related slowing of gait speed may help to prevent or delay the onset of these detrimental health outcomes.

Decreasing energy availability and increasing fatigue may be an underlying driver of age-related declines in gait speed.[12,13] Specifically, lower peak oxygen consumption (VO_2_peak) and reaching fatigue ceilings with less physical activity may lead to compensations, including adaptations of physical behaviors among older adults, such as slowing gait speed.[13][14] Furthermore, previous work suggests that energy requirements during walking and greater perceived physical fatigability, defined as one’s vulnerability to whole-body fatigue anchored to specific tasks, contribute to the development of mobility-related limitations in older adults.[15–19] However, the inter-relationship of walking energetics, broadly defined here as the energetic cost (oxygen consumption) associated with walking at various speeds and intensities, and perceived physical fatigability has not been well-studied. To our knowledge, only one longitudinal study found that those with poorer walking energetics had increased risk of developing greater perceived fatigability.[20] Because both may be risk factors to age-related mobility limitations, it is crucial to understand the role each play with the slowing of gait speed. A more complete understanding may inform the development of lifestyle or therapeutic targets for intervention.

The Study of Energy and Aging-Pilot (SEA-Pilot) found significant differences between slower and faster walkers across multiple walking energetics and fatigue measures, but had limited generalizability due to small sample size (N=36) of primarily high functioning older adults (mean gait speed=1.2 m/s).[15] We extended this work in a large cohort of older adults with a wide range of physical function and fitness to examine the associations of walking energetics and perceived physical fatigability with gait speed and to evaluate whether these associations differed by slower vs faster walkers. We hypothesized that walking energetics, namely lower VO_2_peak, greater energetic cost of walking (EC_w_) during preferred (PWS) and slow walking speed (SWS), and greater cost-capacity ratios (VO_2_ from walking task/VO_2_peak, %), as well as greater perceived physical fatigability are associated with slower gait speed and that the magnitude of these relations are stronger among slower walkers compared to faster walkers. We planned to examine all potential statistical mediation pathways (perceived physical fatigability→walking energetics→gait speed AND walking energetics→perceived physical fatigability→gait speed) to gain a complete understanding of the inter-relationships. However, we hypothesized that the associations of walking energetics with gait speed are statistically mediated by greater perceived physical fatigability as declining energetic reserves may increase fatigue levels among older adults, and that the percent explained is stronger among slower walkers as they are at higher risk of age-related mobility limitations.

## Methods

### Study Sample

The Study of Muscle, Mobility and Aging (SOMMA) (XXXXXXX) is a multi-site prospective cohort study investigating the biological processes associated with aging and has been described elsewhere.[21] Briefly, 879 participants were recruited from two sites (XXXXXXX) and followed longitudinally. The baseline cohort consisted of community-dwelling older men and women aged 70 years and older with body mass index (BMI) ≤40 kg/m^2^ who did not have mobility disability and agreed to undergo muscle tissue biopsy and magnetic resonance scans. Exclusions included self-reported inability to walk one-quarter mile or climb a flight of stairs, active malignancy or dementia, or medical contraindication to biopsy or magnetic resonance scan. Participants must also have been able to complete the 400m walk. During screening, participants who did not appear to be able to complete a 400m walk were asked to complete a 4m walk and were excluded if 4m gait speed was <0.6 m/s. The eligible sample for our analyses includes 849 participants who had no missing data on 4m gait speed, a completed Pittsburgh Fatigability Scale, and at least one walking energetics measure (Figure 1) for final analytic samples. The WIRB-Copernicus Group (WCG) Institutional Review Board (WCGIRB, study number 20180764) approved the study as the single IRB and all participants gave informed written consent.

**Figure 1.**
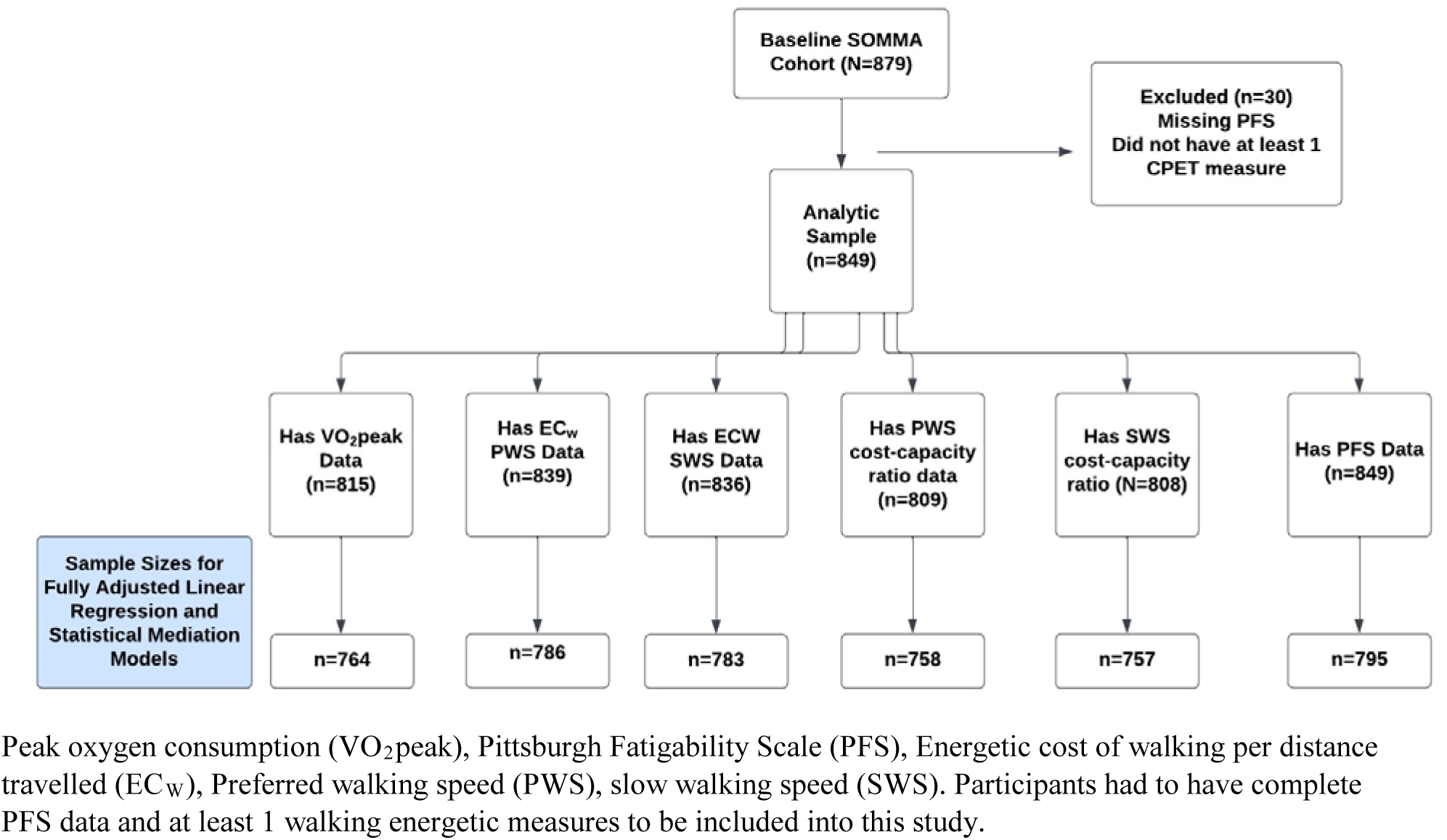
Flowchart of Analytic Sample for Linear Regressions and Statistical Mediation Analyses: The Study of Muscle, Mobility and Aging (SOMMA)

### Walking Energetics Measures

Participants completed a three-phase treadmill cardiopulmonary exercise test (CPET) to collect oxygen consumption (VO_2_, mL/kg/min) across multiple walking intensities. VO_2_ was measured breath-by-breath using a face mask and cardiopulmonary metabolic cart (Medgraphics Ultima Series, Medgraphics Corporation, St. Paul, MN). During Phase 1, participants completed a 5-minute, 0% grade treadmill walk at their PWS, determined from the usual-paced 400m long distance corridor walk speed. Immediately after completion of Phase 1, participants cleared for maximal CPET underwent Phase 2, a symptom-limited maximal modified Balke protocol, in which speed and grade were increased as necessary. Participants were encouraged to reach a respiratory exchange ratio ≥1.05 and/or a Borg Rating of Perceived Exertion (RPE) ≥17 before test termination.[22] VO_2_peak (mL/kg/min) was the highest 30-second average VO_2_ over the course of the test. Participants were given a 20-minute seated rest after completion of Phase 2. Participants not cleared to complete Phase 2 (n=27) were given a 10-minute seated rest before beginning Phase 3. During Phase 3, participants completed a 5-minute, 0% grade slow speed (SWS) treadmill walk. A slow walking speed of 0.67 m/s was used to mimic the minimum walking speed needed for ambulation of daily living.

Average VO_2_ (mL/kg/min) from Phase 1 and 3 walks were calculated from the last 3 minutes of each test. Phase 1 and 3 VO_2_ were also divided by walking speed to determine the energetic cost of walking (EC_W_) per distance travelled (mL/kg/meter) at PWS and SWS. Two cost-capacity ratios of VO_2_ from Phase 1 (PWS) and 3 (SWS) relative to VO_2_peak were calculated (%, VO_2_/VO_2_peak) to understand relative intensity (PWS) and the capacity needed to maintain ambulation (SWS, higher ratio=higher energetic cost for task).

### Perceived Physical Fatigability

Perceived physical fatigability was measured using the validated Pittsburgh Fatigability Scale (PFS).[23,24] The PFS is a 10-item scale that asks participants to rate the level of physical and mental fatigue they expected or imagined they would feel after performing each task (0=no fatigue, 5=extreme fatigue). Each item implied a specific intensity and duration (e.g. leisurely walk for 30 minutes). Participants were asked to respond to all items regardless of whether they have performed that activity in the past month. Each subscale score ranged from 0 to 50; a higher score indicates greater perceived fatigability. Scores were imputed for participants who did not respond to ≤3 items within the scale using established methods (n=18).[25] We only examined PFS Physical score for this analysis.

### 4m Gait Speed

Gait speed was calculated using the faster of two usual-paced 4m trials. Because of the clinical utility of the 4m gait speed test and high correlation with gait speed calculated from longer walking-based tasks (r=0.74 between 4m and 400m walks),[26] we chose this measure as our primary outcome. Participants were categorized as slower (<1.01 m/s) vs faster (≥1.01 m/s) walkers using median 4m gait speed.

### Covariates

Age, sex, race/ethnicity (white vs nonwhite), and smoking status were self-reported. Height without shoes and weight with light clothing were measured and used to calculate BMI (kg/m^2^). We queried self-report of a physician diagnosis history (yes/no) of several health conditions. A composite multimorbidity index was calculated using a modified list of chronic conditions from the Rochester Epidemiology Project.[27] Physical activity was objectively measured using a 3-axial accelerometer (ActiGraph GT9X) worn on the non-dominant wrist, with a goal of collecting at least 7 consecutive 24-hour periods (valid wear=≥17 hours wear during 24-hour period). Daily activity was assessed as averaged total activity count per 24-hour valid day.

### Statistical Analysis

Descriptive characteristics of participants were reported (mean±standard deviation (SD) or frequencies). Comparisons by slower versus faster walking status were evaluated using t-tests for normally distributed continuous variables, Wilcoxon rank-sum test for skewed continuous variables, and for categorical variables a chi-square test or a Fisher’s exact test for those with low expected cell counts. We used linear regressions to examine associations of walking energetics and perceived fatigability with 4m gait speed. We also stratified linear regressions by slower and faster walkers to examine whether associations were differential.

We used a statistical mediation approach (PROC CAUSALMED) to examine the percent explained by perceived physical fatigability on the association between walking energetics and gait speed overall, and by slower and faster walkers. This non-parametric approach specifies the direct and indirect effects of the independent variable on the dependent variable and the mediation percentage.[28] Higher mediation percentage indicates the mediator variable is explaining a greater proportion of the association of the independent variable on the dependent variable. We first examined whether perceived physical fatigability was a statistical mediator between walking energetics and gait speed. Next, we examined the alternative pathway (perceived physical fatigability→walking energetics-→gait speed) for complete understanding of the inter-relationships. All models were adjusted for age, sex, race, total physical activity counts, and clinic site. All analyses were generated using SAS software version 9.4 (SAS Institute Inc, Cary, NC).

## Results

The full sample of 849 participants (58.7% Women, 85.9% White, 76.3±4.9 years old) had a mean 4m gait speed of 1.04±0.20 m/s and mean VO_2_peak of 20.2±4.8mL/kg/min. Mean PFS Physical score was 15.7±8.6, with 53.5% of participants classified as having more severe perceived physical fatigability (PFS Physical score ≥15). Compared to faster walkers (≥1.01 m/s), slower walkers (<1.01m/s) on average were older, more likely to be women, less likely to be White, had higher BMI, had more comorbidities and lower physical activity (p<0.05, Table 1). Slower walkers had nearly 4 mL/kg/min lower VO_2_peak (18.3 vs 22.0 mL/kg/min, p<0.0001), higher EC_w_ PWS (0.21 vs 0.19, p<0.001), and 9% higher SWS cost-capacity ratio (53.5% vs 44.4%, p<0.0001). Slower walkers also had nearly a 6-point higher PFS Physical score (18.2 vs 13.3 points, p<0.0001) (Figure 2).

**Figure 2.**
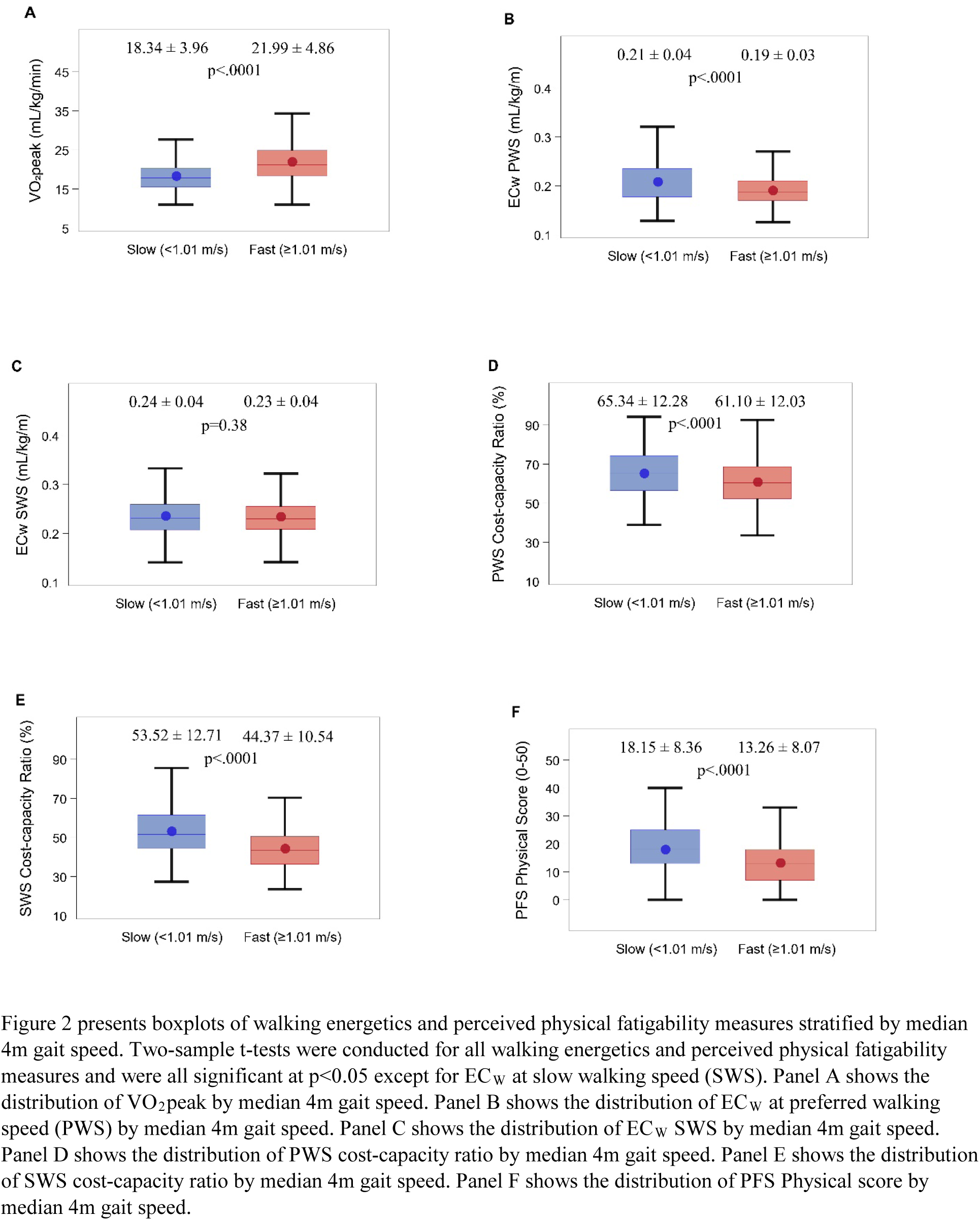
Comparisons of Walking Energetics and Perceived Physical Fatigability, stratified by Slower vs Faster Walkers: The Study of Muscle, Mobility and Aging.

**Table 1.** Descriptive Characteristics of the Study of Muscle, Mobility and Aging (SOMMA) Participants (N=849), by Median 4m Gait Speed.

| | Overall | Slower Walkers<br>($<1.01$ m/s)<br>N=416 | Fast Walkers<br>($\geq 1.01$ m/s)<br>N=433 | p-value |
| --- | --- | --- | --- | --- |
| Age, years | 76.3 ± 5.0 | 77.5 ± 5.6 | 75.2 ± 4.0 | <.0001 |
| Women | 498 (58.7) | 261 (62.7) | 237 (54.7) | 0.01 |
| White | 729 (85.9) | 341 (82.0) | 388 (89.6) | 0.001 |
| Body mass index, kg/m<sup>2</sup> | 27.5 ± 4.6 | 28.5 ± 4.7 | 26.6 ± 4.2 | <.0001 |
| SOMMA multimorbidity index (0-11)<sup>†</sup> | 0.8 ± 0.9 | 0.9 ± 0.9 | 0.7 ± 0.8 | 0.02 |
| 1. Cancer* | 212 (25.0) | 103 (24.8) | 109 (25.2) | 0.8 |
| 2. Cardiac arrhythmia | 34 (4.0) | 23 (5.5) | 11 (2.5) | 0.02 |
| 3. Chronic kidney disease or renal failure | 30 (3.5) | 17 (4.1) | 13 (3.0) | 0.3 |
| 4. Chronic obstructive pulmonary disease or other lung disease | 111 (13.1) | 54 (13.0) | 57 (13.2) | 0.9 |
| 5. Coronary artery disease | 57 (6.7) | 31 (7.5) | 26 (6.0) | 0.3 |
| 6. Congestive heart failure | 6 (0.7) | 2 (0.5) | 4 (0.9) | 0.6 |
| 7. Dementia | 0 | 0 | 0 |  |
| 8. Depression | 70 (8.3) | 42 (10.2) | 28 (6.5) | 0.05 |
| 9. Diabetes mellitus | 126 (14.8) | 73 (17.5) | 53 (12.2) | 0.02 |
| 10. Stroke | 21 (2.5) | 11 (2.6) | 10 (2.3) | 0.7 |
| 11. Aortic stenosis | 8 (0.9) | 4 (1.0) | 4 (0.9) | 1.0 |
| Smoking Status |  |  |  |  |
| Never | 479 (56.6) | 227 (54.8) | 252 (58.2) | 0.3 |
| Past | 343 (40.5) | 172 (41.6) | 171 (39.5) |  |
| Current | 25 (3.0) | 15 (3.6) | 10 (2.3) |  |
| Total Activity Counts per 100,000 counts | 19.9 ± 5.9 | 19.2 ± 5.9 | 20.5 ± 5.8 | 0.001 |
| 4m Gait Speed, m/s | 1.04 ± 0.2 | 0.88 ± 0.09 | 1.20 ± 0.16 | <0.001 |

Note: mean ± standard deviation or n (%)
<sup>†</sup>The SOMMA multimorbidity index score included 11 age-related conditions: cancer, chronic kidney disease or renal failure, atrial fibrillation, lung disease (i.e., chronic obstructive pulmonary disease, bronchitis, asthma, or emphysema), coronary heart disease (i.e., blocked artery or myocardial infarction), depression [(e.g., $\geq 10$ score on the 10-item Center for Epidemiologic Studies Depression Scale (CESD-10)], heart failure, dementia, diabetes, stroke, and aortic stenosis
\*excludes nonmelanoma skin cancer

### Associations of Walking Energetics and Perceived Physical Fatigability on 4m Gait Speed

Each SD higher increment in fitness (VO_2_peak) was associated with faster walking speeds of 0.1 m/s after adjustment. Each SD higher increment in worse efficiency while walking (EC_W_ PWS, PWS and SWS cost-capacity ratio) was associated with slower walking speeds of 0.02-0.09 m/s after adjustment (Table 2). Associations for VO_2_peak, and SWS cost-capacity ratio were stronger among faster walkers compared to slower walkers; associations were not different between slower and faster walkers for EC_W_ PWS or PWS cost-capacity ratio (Table 2). There were no significant associations between EC_W_ SWS and 4m gait speed.

**Table 2.** Associations of Walking Energetics and Perceived Fatigability on 4m Gait Speed: The Study of Muscle, Mobility and Aging (SOMMA)

|  | Per<br>increment | Overall | Slower Walkers | Faster Walkers |
| --- | --- | --- | --- | --- |
| | SD | $\beta$ (95% CI) | $\beta$ (95% CI) | $\beta$ (95% CI) |
| **Walking Energetics** |  |  |  |  |
| VO<sub>2</sub>peak (mL/kg/min) | 4.81 | 0.10 (0.08, 0.12)* | 0.02 (0.01, 0.04)* | 0.07 (0.05, 0.09)* |
| EC<sub>w</sub> at PWS (mL/kg/m) | 0.04 | -0.05 (-0.07, -0.04)* | -0.02 (-0.03, -0.01)* | -0.03 (-0.04, -0.01)* |
| EC<sub>w</sub> at SWS (mL/kg/m) | 0.04 | -0.01 (-0.03, 0.003) | -0.01 (-0.01, 0.00) | 0.00 (-0.02, 0.02) |
| Cost-capacity Ratio<br>(VO<sub>2</sub> PWS / VO<sub>2</sub>peak) | 12.33 | -0.02 (-0.03, -0.004)* | -0.01 (-0.02, 0.00) | -0.01 (-0.03, 0.01) |
| Cost-capacity Ratio<br>(VO<sub>2</sub> SWS / VO<sub>2</sub>peak) | 12.49 | -0.09 (-0.10, -0.07)* | -0.02 (-0.03, -0.01)* | -0.05 (-0.08, -0.04)* |
| **Perceived Physical Fatigability** |  |  |  |  |
| PFS Physical score, 0-50 | 8.57 | -0.06 (-0.07, -0.04)* | -0.02 (-0.03, -0.01)* | -0.03 (-0.05, -0.01)* |

Each row indicates a separate linear regression model with 4m gait speed as the outcome, adjusted for clinic site, age, race (white vs nonwhite), sex, and mean sum of total physical activity counts, measured by ActiGraph GT9X accelerometer
Results are presented per 1 SD increment for explanatory variable. The SD for the overall group was used in the stratified results.
\*Indicates significant p-values at the $\leq 0.05$ level
EC<sub>w</sub> = Energetic cost of walking; PWS = Preferred walking speed; SWS = Slow walking speed; PFS = Pittsburgh Fatigability Scale

A 1SD higher increment in PFS Physical score was associated with a 0.06 m/s (95% CI:- 0.07, -0.04) slower gait speed after adjustment. Associations did not differ between slower and faster walkers (Table 2).

### Examining PFS Physical Score as a Mediator Between Walking Energetics and Gait Speed

The mediation analysis revealed that PFS Physical score explained 15.2% (95% CI:8.7%, 21.8%) of the total association between VO_2_peak and gait speed after adjustment. Among slower walkers, PFS Physical score explained 34.2% (95% CI:9.7%, 58.6%) of the total association, whereas it only explained 4.6% (95% CI:-4.1,13.3%) of the total association among faster walkers. When examining the association between SWS cost-capacity ratio and gait speed, PFS Physical Score explained 15.9% (95% CI:9.3%, 22.5%) of the total associations. Similar to VO_2_peak model, the mediation percentage of PFS Physical score on the association of SWS cost-capacity on gait speed was larger among slower walkers (17.9%; 95% CI:5.2%, 30.6%) compared to faster walkers (9.0%; 95% CI: -2.9%, 20.9%) (Figure 2). The percent explained by PFS Physical score between EC_W_ PWS and gait speed was 10.7% (95% CI:3.6%, 17.9%) (Figure A3).

**Figure 3.**
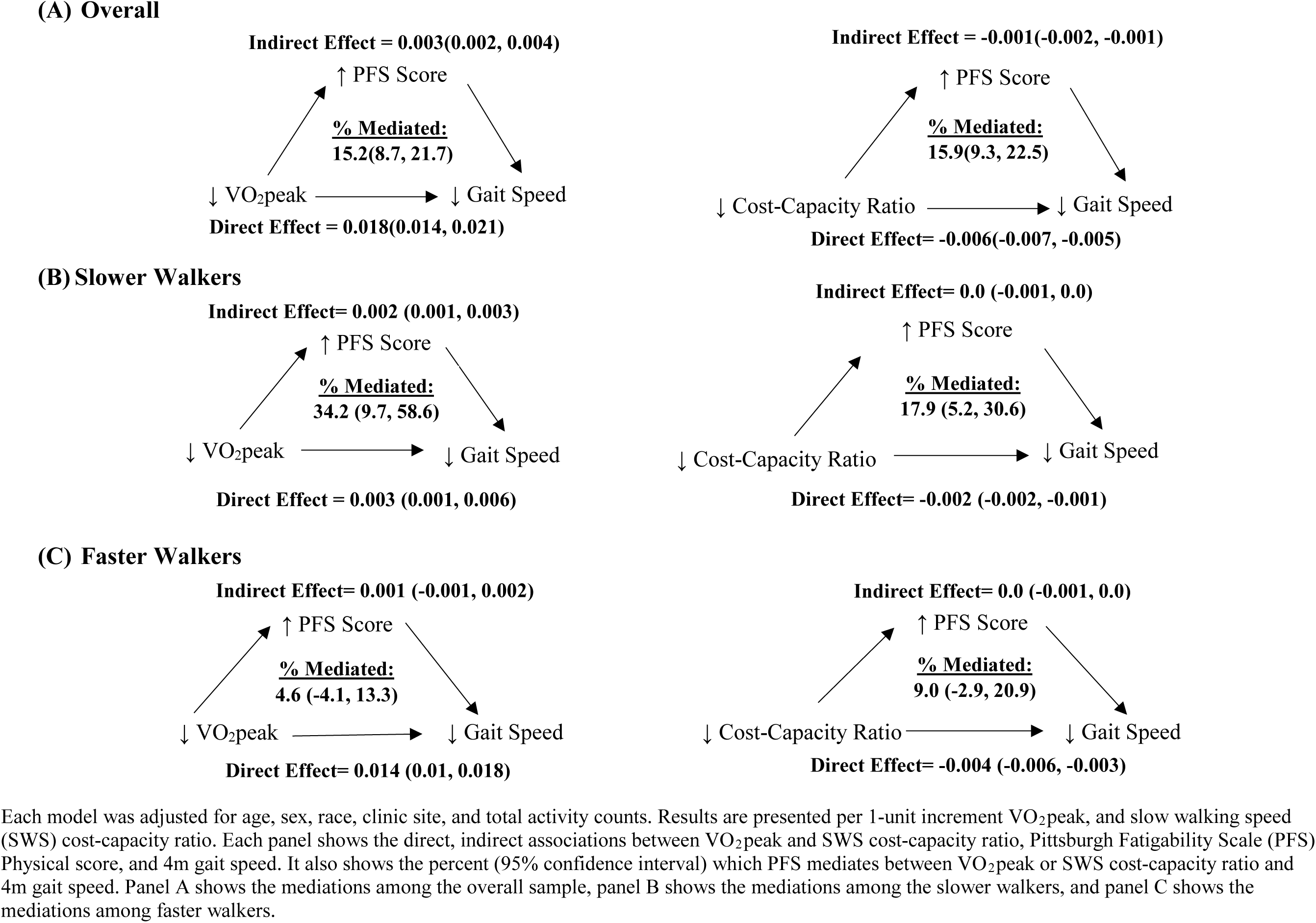
Examining Perceived Physical Fatigability as a Mediator between Walking Energetics and 4m Gait Speed: The Study of Muscle, Mobility and Aging (SOMMA)

### Understanding Walking Energetics as a Mediator between PFS Physical Score and Gait Speed

VO_2_peak, SWS cost-capacity ratio, and EC_W_ at PWS explained a larger proportion of the total association between PFS Physical score and gait speed (VO_2_peak: 39.3% (95% CI:27.2%, 51.4%); SWS cost-capacity Ratio: 31.5%, (95% CI:20.8%, 42.2%); EC_W_ PWS: 8.8% (95% CI:2.8%, 14.85%). Mediation percentages were higher among faster walkers compared to slower walkers (Appendix, Figures A1 and A2). VO_2_peak explained 66.2% (95% CI:22%, 110.4%) vs 12.7% (95% CI:0.7%, 24.7%) and SWS cost-capacity ratio explained 48.0% (95% CI:11.8%, 84.3%) vs 15.4% (95% CI:4.1%, 26.7%) among faster vs slower walkers.

## Discussion

In a sample of community-dwelling older men and women, we observed significant associations of walking energetics and perceived physical fatigability with gait speed. Additionally, our statistical mediation results suggested that perceived physical fatigability may be an important contributor to gait speed among slower walkers, whereas walking energetics, specifically VO_2_peak and SWS cost-capacity ratio, may largely contribute to gait speed among faster walkers.

Our finding of significant associations between walking energetics and gait speed in SOMMA confirms and extends previous work. Specifically, reports from the Baltimore Longitudinal Study of Aging (BLSA) show an association between EC_W_ PWS, VO_2_peak, and cost-capacity ratios with slow gait speed.[16,18,19] Similarly, the SEA-Pilot found significant associations between VO_2_peak and PWS VO_2_ with slow gait speed.[15] Preferred walking speeds tend to be chosen to minimize energetic cost of walking.[29] This trend is consistent with aging, despite older adults having slower gait speed and higher energetic costs of walking compared to younger adults.[30] While we found that EC_w_ PWS was significantly associated with gait speed, VO_2_peak and SWS cost-capacity ratio, both measures of functional capacity, were more strongly associated with gait speed in the SOMMA cohort and these associations were stronger among faster walkers compared to slower walkers. When functional capacity is high, older adults may exhibit fewer physiological symptoms of exertion (e.g. heart rate) while walking at higher intensity and speed and thus lower perceptions of fatigue. Recently, mitochondrial dysfunction has been proposed as the primary mechanism of slow walking among older adults.[31,32] Mitochondrial dysfunction is associated with both VO_2_peak and perceived physical fatigability,[33–35] suggesting that mitochondrial function may be an important upstream cause of limitations due to fitness and fatigue and should be further explored.

Interestingly, SWS cost-capacity ratio was associated with slower gait speed at a magnitude similar to that of VO_2_peak. SWS cost-capacity ratios >50% indicate greater risk of mobility limitations.[16] In SOMMA, the average SWS cost-capacity ratio among the overall sample was 48%, but was 53.5% among slower walkers, indicating that those with slower gait speeds (<1.01 m/s) may be at risk for mobility limitation. Recent work in BLSA has shown that cost-capacity ratio increases accelerates with age.[36] Further work understanding longitudinal associations between cost-capacity ratio and gait speed are needed to understand the impact of each other during aging.

Previous literature shows an association between greater perceived physical fatigability and slow gait speeds.[15,37–39] The PFS is a highly sensitive marker of impending functional decline.[38] Our results suggest that periodic clinical assessment of perceived physical fatigability using the PFS may be warranted. We did not include RPE fatigability measured during slow walking because the slow walk may not be strenuous enough to elicit perceptions of exertion among faster walkers.

Since this is a cross-sectional study, we were not able to make any causal inference or infer temporality. However, our statistical mediation analyses provided meaningful insights on the associations of fitness and fatigability on gait speed. Lower energetic reserves are associated with risk of developing greater perceived physical fatigability,[20] suggesting that fatigability may mediate the associations between walking energetics and gait speed. Our results support this as PFS Physical score was a significant mediator between walking energetic measures and gait speed. Interestingly, differences in mediation pathways were observed between slower and faster walkers, with PFS Physical score explaining a third of the association between VO_2_peak and gait speed among slower walkers, but only 5% among faster walkers. Greater perceived physical fatigability may be a more downstream marker of mobility decline compared to walking energetics. Those who walk at or below 1.0 m/s have lower fitness and poorer energetic measures. As a result of lower fitness and energetics, these older adults may have already experienced some functional decline and may be primarily limited by their fatigue thresholds. Conversely, older adults who walk above 1.0 m/s may not have experienced meaningful declines in fitness and therefore may not be limited by fatigue. Our examination of the alternative statistical mediation pathway highlights that both perceptual and physiological indices influence gait speed and should be studied in tandem.

Based on our results, it seems logical that future interventions targeting mobility decline should integrate cardiorespiratory fitness, walking and energy use efficiency, and target perceptions of fatigue. Furthermore, interventions can be enhanced if tailored to baseline gait speed. Among faster walkers, interventions are likely to be most effective if focused on improving and maintaining physical fitness. Among slower walkers, interventions are likely to be most effective if focused on reducing perceived fatigability, possibly through a physical activity intervention.[40] Additional research is indicated.

Our study has some limitations. First, the cross-sectional design does not allow for establishment of temporality or causal inference. As such, it is still unknown whether walking energetics or perceived physical fatigability come first in the mobility decline causal pathway. Second, while SOMMA’s race/ethnicity distributions reflect the communities in which they were recruited, the cohort is not diverse, limiting the generalizability. Finally, walking energetics was measured on a treadmill. Overground walking may be preferred to treadmill walking to obtain more real-world measures of VO_2_ and EC_W_. Despite these limitations, this study has many strengths. All walking energetics were measured using gold standard CPET, which allowed for objective and robust analyses of walking efficiency and effort. Additionally, perceived physical fatigability was measured using a well-established, validated method. Finally, the SOMMA cohort includes a wide range of physical function and fitness.

In conclusion, our study revealed that walking energetics and perceived physical fatigability are associated with gait speed. Additionally, differences existed in mediation between slower and faster walkers, with greater perceived fatigability acting as the primary statistical mediator among slower walkers and walking energetics among faster walkers. Thus, the driving factors of mobility decline may vary during progression in the age-related disablement pathway. Longitudinal work is needed to establish the causal pathway of mobility decline as well as to better understand the associations between walking energetics and perceived physical fatigability among older adults.

## Data Availability

All data produced in the present study are available upon reasonable request to the authors

https://sommaonline.ucsf.edu

## Acknowledgements

Preliminary data were presented at the American College of Sports Medicine Annual Meeting, Denver, CO on May 31, 2023.

## Appendix for “Role of Walking Energetics and Perceived Fatigability on Mobility Differ by Walking Speed: Findings from the Study of Muscle, Mobility and Aging (SOMMA)”

**Figure A1.**
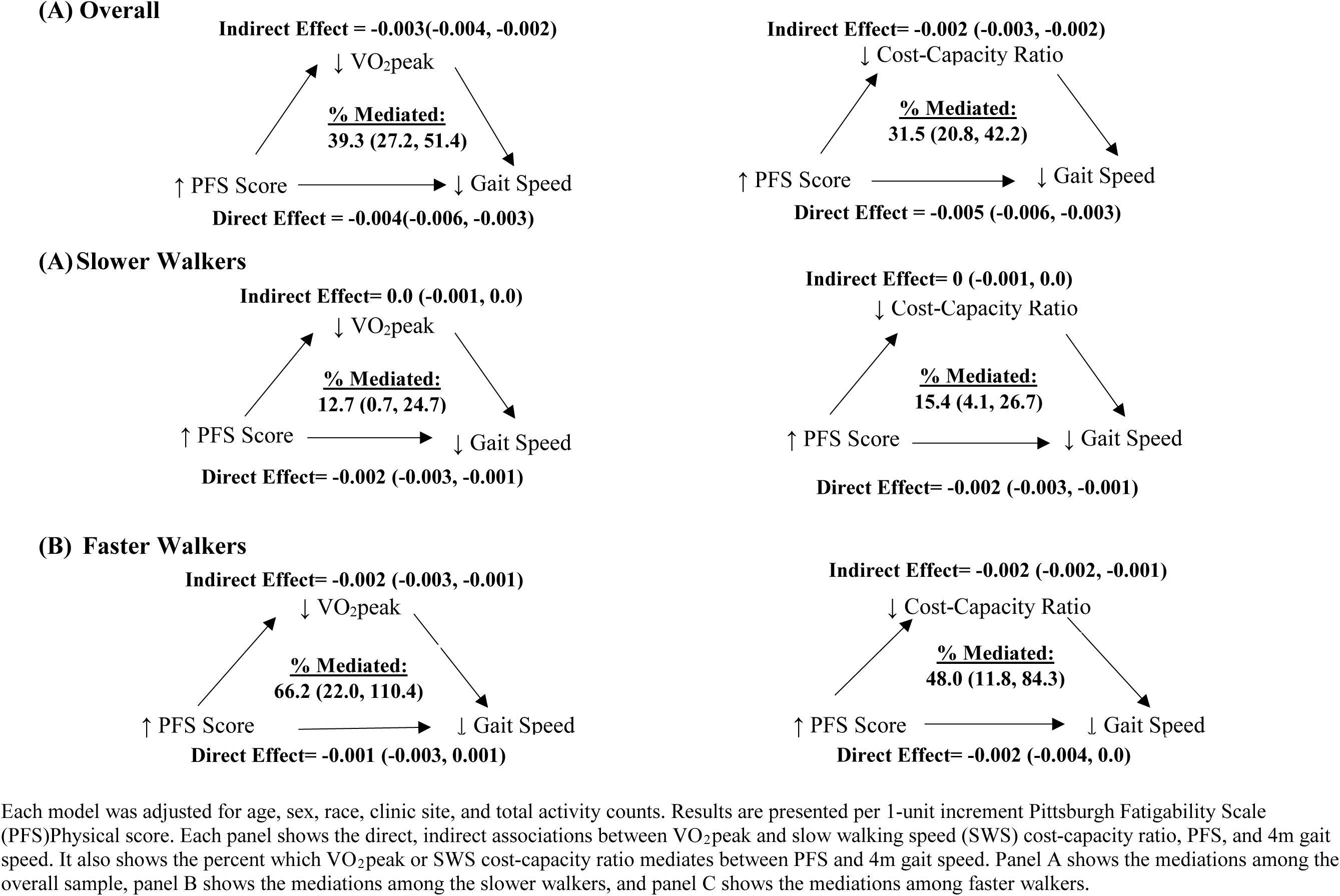
Examining Walking Energetics as a Mediator between Perceived Physical Fatigability and 4m Gait Speed: The Study of Muscle, Mobility and Aging (SOMMA)

**Figure A2.**
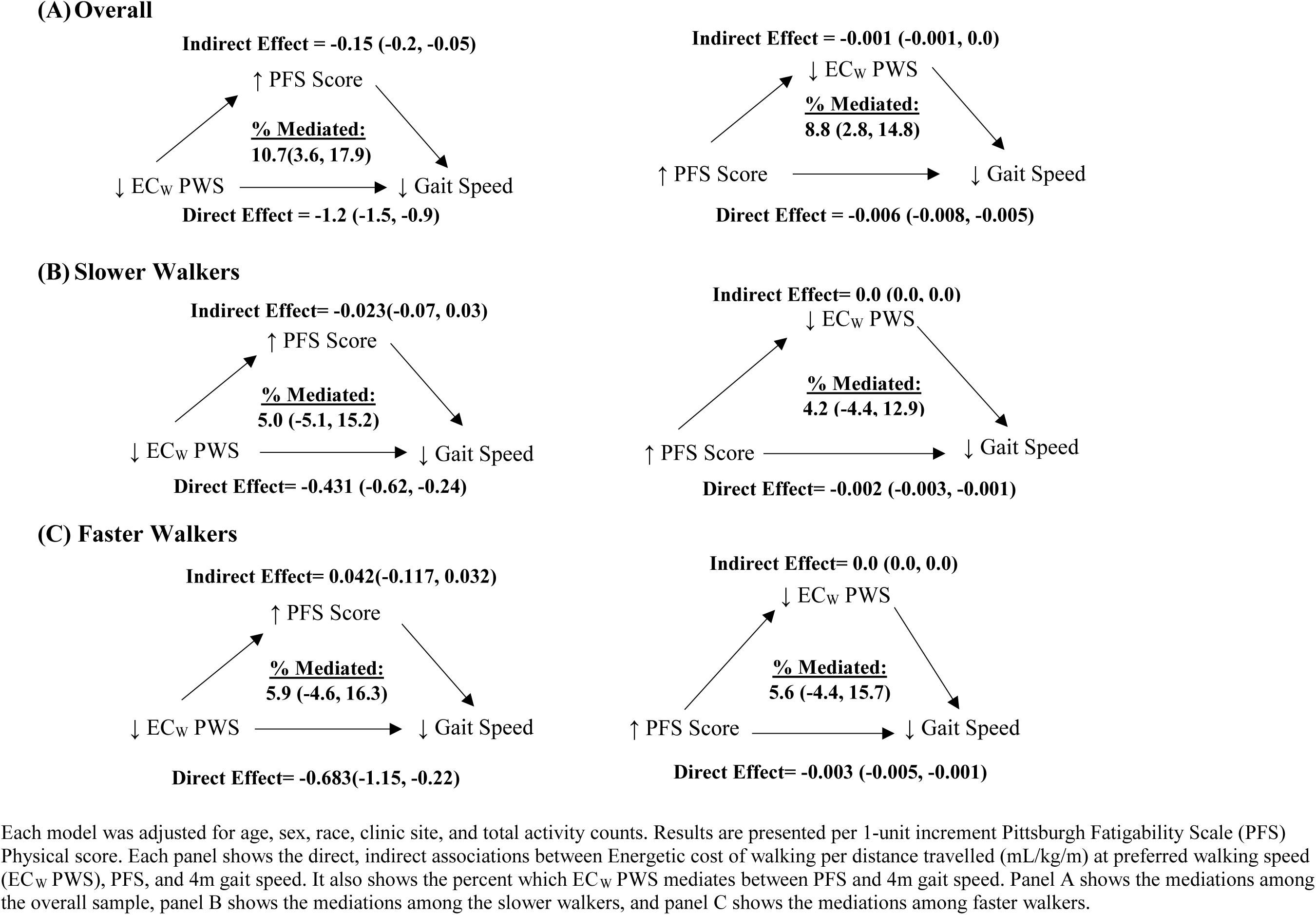
Examining the Mediations of Perceived Physical Fatigability and Energetic Cost of Walking at Preferred Walking Speed with 4m Gait Speed: The Study of Muscle, Mobility and Aging (SOMMA)

## Notes

**Declaration of Sources of Funding:** This work was supported by funding from the National Institute on Aging (AG 059416). Study infrastructure support was funded in part by NIA Claude D. Pepper Older American Independence Centers at University of Pittsburgh (P30 AG024827) and Wake Forest University (P30 AG021332) and the Clinical and Translational Science Institutes, funded by the National Center for Advancing Translational Science, at Wake Forest University (UL1 0TR001420). Additionally, the Claude D. Pepper Older Americans Independence Center, Research Registry and Developmental Pilot Grant (NIH P30 AG024827), and the Intramural Research Program, National Institute on Aging supported N.W.G to develop the Pittsburgh Fatigability Scale.

### Competing Interest Statement

The authors have declared no competing interest.

### Funding Statement

This work was supported by funding from the National Institute on Aging (AG 059416). Study infrastructure support was funded in part by NIA Claude D. Pepper Older American Independence Centers at University of Pittsburgh (P30 AG024827) and Wake Forest University (P30 AG021332) and the Clinical and Translational Science Institutes, funded by the National Center for Advancing Translational Science, at Wake Forest University (UL1 0TR001420). Additionally, the Claude D. Pepper Older Americans Independence Center, Research Registry and Developmental Pilot Grant (NIH P30 AG024827), and the Intramural Research Program, National Institute on Aging supported N.W.G to develop the Pittsburgh Fatigability Scale.

### Author Declarations

The WIRB-Copernicus Group (WCG) Institutional Review Board (WCGIRB, study number 20180764) gave ethical approval for this study as the single IRB and all participants gave informed written consent.

